# Predicting the efficacy of exenatide in Parkinson’s disease using genetics – a Mendelian randomization study

**DOI:** 10.1101/2020.10.20.20215855

**Authors:** Catherine S. Storm, Demis A. Kia, Mona Almramhi, Dilan Athauda, International Parkinson’s Disease Genomics Consortium (IPDGC), Stephen Burgess, Thomas Foltynie, Nicholas W. Wood

## Abstract

**Background:** Exenatide is a glucagon-like peptide 1 receptor (GLP1R) agonist used in type 2 diabetes mellitus that has shown promise for Parkinson’s disease in a phase II clinical trial. Drugs with genetic evidence are more likely to be successful in clinical trials. In this study we investigated whether the genetic technique Mendelian randomization (MR) can “rediscover” the effects of exenatide on diabetes and weight, and predict its efficacy for Parkinson’s disease.

**Methods:** We used genetic variants associated with increased expression of *GLP1R* in blood to proxy exenatide, as well as variants associated with expression of *DPP4, TLR4* and 15 genes thought to act downstream of GLP1R or mimicking alternative actions of GLP-1 in blood and brain tissue. Using an MR approach, we predict the effect of exenatide on type 2 diabetes risk, body mass index (BMI), Parkinson’s disease risk and several Parkinson’s disease progression markers.

**Results:** We found that genetically-raised *GLP1R* expression in blood was associated with lower BMI and possibly type 2 diabetes mellitus risk, but not Parkinson’s disease risk, age at onset or progression. Reduced *DPP4* expression in brain tissue was significantly associated with increased Parkinson’s disease risk.

**Conclusions:** We demonstrate the usefulness of MR using expression data in predicting the efficacy of a drug and exploring its mechanism of action. Our data suggest that GLP-1 mimetics like exenatide, if ultimately proven to be effective in Parkinson’s disease, will be through a mechanism that is independent of GLP1R in blood.

## Introduction

Modern drug development is remarkably costly and time consuming. It takes approximately $1.3 billion and over a decade for a drug to proceed from initial testing in humans to licensing (Wouters, McKee, and Luyten 2020), and 90% of drugs that enter phase I clinical trials never proceed to be launched (Smietana, Siatkowski, and Møller 2016). Insufficient safety or efficacy are the most common reasons drug development projects are unsuccessful, and medications for central nervous system disorders are particularly likely to fail (Kesselheim, Hwang, and Franklin 2015). One strategy that circumvents safety problems is drug repurposing, where already-licensed drugs are used for new medical indications. Since licenced medications have passed safety assessment in humans, the same toxicology studies do not need to be repeated and so these drugs could reach patients both sooner and at a much lower cost (Pushpakom *et al*., 2018). There are major patent- and regulatory barriers to drug repurposing, and robust demonstration of efficacy is a critical step in creating an incentive to invest (Pushpakom *et al*., 2018). As such, more accurate and cost-effective approaches for drug target validation must be found.

Drugs with genetic evidence are considerably more likely to be efficacious (Nelson et al. 2015), and Mendelian randomization (MR) is a genetic technique that can obtain human evidence for efficacy early in the drug development pipeline. MR builds on the principle that genetic variants associated with an environmental risk factor mimic exposure thereto (Hemani et al. 2018; Evans and Davey Smith 2015). For example, a genetic propensity for lower blood glucose is similar to receiving a low-dose glucose-lowering drug throughout life. Similarly, genetic variants that are associated with reduced expression levels of a gene (expression quantitative trait loci, eQTLs) can be used as proxies to mimic a pharmacological antagonist of the encoded proteins (Storm et al. 2020; Schmidt et al. 2020).

Parkinson’s is a neurodegenerative movement disorder for which finding disease-modifying treatments has been a great challenge. In recent years, the drug exenatide has shown promise in a phase II clinical trial for Parkinson’s (Athauda et al. 2017). Exenatide is a medication used to treat type 2 diabetes mellitus, and it is also known to cause weight loss. As a glucagon-like peptide 1 mimetic, exenatide is thought to act on the GLP-1 receptor (GLP1R). The protein DPP-4 breaks down GLP-1 *in vivo*, and there is evidence that toll-like receptor 4 (TLR4) may be necessary for intestinal GLP-1 secretion in mice (Wang et al. 2019).

In this study we assessed whether MR and eQTL data for the GLP1R pathway can (1) “rediscover” the use of exenatide as a treatment for type 2 diabetes mellitus and its effect on weight. We then extend this tool to (2) predict the likely efficacy of this drug for Parkinson’s.

## Methods

MR analyses were performed using R software version 3.6.1 (R Core Team 2019) with the R packages “TwoSampleMR” (Hemani et al. 2018) and “MendelianRandomization” (Yavorska and Burgess 2017). All expression and GWAS data used are openly available, and full details about the recruitment and analyses are provided in the original publications.

### Mimicking exenatide - genetic instrument development

We used SNPs associated with the expression of the *GLP1R, DPP4* and *TLR4* genes in blood provided by the eQTLGen consortium (blood samples from 31 684 mostly European-ancestry individuals). For Parkinson’s-related outcomes, we also looked at gene expression data from brain tissue, available from the PsychENCODE consortium (brain tissue samples from mostly European-ancestry individuals: 679 healthy controls, 497 schizophrenia, 172 bipolar disorder, 31 autism spectrum disorder, 8 affective disorder patients) (Võsa et al. 2018; Wang et al. 2018). We included all SNPs with p < 5 × 10^−5^. In a secondary analysis, we identified SNPs associated with the expression of 15 genes encoding proteins hypothesized to be involved in exenatide’s mechanism of action in Parkinson’s: *AKT1, AKT2, AKT3, FOXO1, FOXO3, GCG, GSK3B, IRS1, MAPK11, MAPK12, MAPK13, MAPK14, MTOR, NFKB1*, and *NFKB2* (Athauda and Foltynie 2016; Athauda et al. 2019).

### Outcome data

Exenatide is a licensed treatment for type 2 diabetes mellitus, and this drug is known to cause weight loss. We therefore used openly available GWAS summary statistics for type 2 diabetes mellitus risk (62 892 cases, 592 424 controls) and body mass index (BMI; ∼700 000 individuals) to ascertain if MR using eQTLs can “rediscover” the known effects of exenatide (Xue et al. 2018; Yengo et al. 2018).

For Parkinson’s, we used data pertaining to: disease risk (15 056 cases, 18 618 proxy cases, 449 056 controls), age at onset (17 996 cases) and 13 markers of progression (4 093 cases): total Unified Parkinson’s Disease Rating Scale (UPDRS)/Movement Disorder Society revised version total (Parkinson’s progression rating scale), UPDRS parts 1 to 4 (1 = non-motor symptoms, 2 = motor symptoms, 3 = motor examination, 4 = motor complications), MOCA (cognitive impairment), MMSE (cognitive impairment), SEADL (activities of daily living and independence), dementia, depression, dyskinesia, Hoehn and Yahr stage (Parkinson’s progression rating scale), and reaching Hoehn and Yahr stage 3 or more (Nalls et al. 2019; Blauwendraat et al. 2019; Iwaki et al. 2019).

### Main MR analysis and quality control

For the main analyses, SNPs were clumped at *r*^2^ = 0.2; this means that if the squared correlation coefficient (*r*^2^) of two eQTLs for the same gene is greater than 0.2, only the eQTL with the smallest p-value will be retained. We applied Steiger filtering to all analyses to remove any genes where SNPs explain a greater proportion of variation in the disease outcome than variation in the exposure (gene expression). A Wald ratio was calculated for each SNP, and for each gene Wald ratios were meta-analysed using inverse-variance weighted (IVW), MR-Egger and maximum likelihood methods, incorporating an LD-matrix to account for correlation for genes where > 2 SNPs were available (Burgess et al. 2015). The MR-Egger intercept, Cochran’s Q and *I*^2^ tests were used to check for directional pleiotropy and heterogeneity between SNPs (Hemani et al. 2018; Yavorska and Burgess 2017). P-values were adjusted for multiple testing using the false discovery rate (FDR) method, correcting for the number of genes tested.

We used the principal-components-based IVW (IVWPC), factor-based limited information maximum likelihood (F-LIML), and factor-based conditional likelihood ratio (F-CLR) methods as secondary analyses to probe the robustness of the *GLP1R*-diabetes association (Patel et al. 2020; Burgess et al. 2017). These methods exploit correlation between SNPs and build new instruments using principal components or factor analysis, as indicated by the name. This is beneficial because highly correlated variants can be included, and there is evidence that especially the F-CLR method is robust regardless of instrument strength (Patel et al. 2020). Here, SNPs were clumped at *r*^2^ = 0.6, which allows for more correlation between eQTLs and so retains a larger number of SNPs per gene (compared to an *r*^2^ cut-off of 0.2). For the IVWPC method, we included principal components explaining 99% of variation in the weighted correlation matrix (Burgess et al. 2017).

## Results

In the main analysis, increased expression of *GLP1R* predicted a reduced diabetes risk at nominal significance; *DPP4* and *TLR4* expression were not associated with type 2 diabetes mellitus risk (Figure 1a and Figure S1; Table S1). Raised *GLP1R* expression predicted a significantly reduced BMI, which is consistent with weight loss seen with exenatide use (Figure 1c and Figure S2; Table S1). *GLP1R* passed the MR-Egger intercept and Cochran’s Q tests for diabetes (MR-Egger intercept *p* = 0.268, Cochran’s Q *p* = 0.452, *I*^2^ = 0) and BMI (MR-Egger intercept *p* = 0.173, Cochran’s Q *p* = 0.107, *I*^2^ = 0.337). We found similar results when using the maximum likelihood method, and the MR-Egger estimate tended in the same direction of effect.

**Figure 1:**
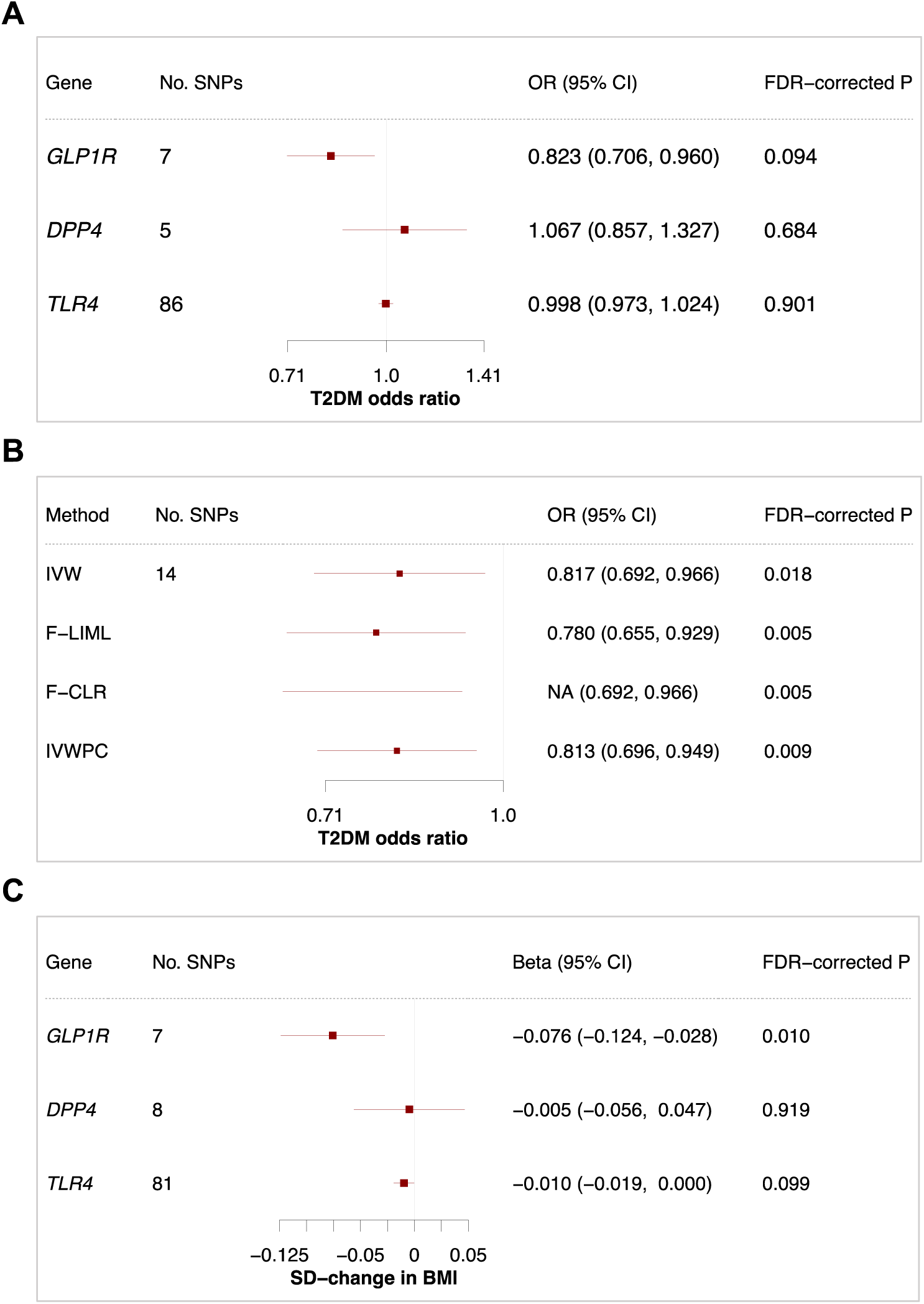
Forest plots illustrating the MR estimates of GLP1R, DPP4 and TLR4 in blood. All results computed per 1-standard-deviation increase in gene expression. P-values were corrected for the number of genes tested using the FDR method. (A) Wald ratio or IVW estimates of GLP1R, DPP4 and TLR4 in blood for type 2 diabetes mellitus, clumping at r^2^ = 0.2. (B) Results for GLP1R in type 2 diabetes mellitus using IVW, F-LIML, F-CLR and IVWPC methods, clumping at r^2^ = 0.6. The F-CLR method provides a confidence interval and p-value, but not a point estimate. (C) Wald ratio or IVW estimates of GLP1R, DPP4 and TLR4 in blood for BMI, clumping at r^2^ = 0.2. 95% CI, 95% confidence interval; NA, not applicable; OR, odds ratio; SD, standard deviation; T2DM, type 2 diabetes mellitus.

Since exenatide is a known drug for diabetes mellitus, we were surprised to find that this effect did not remain significant upon multiple testing. Many SNPs are lost during clumping at *r*^2^ = 0.2, so we repeated the analysis using the IVWPCA and F-CLR methods, which exploit linkage between SNPs and therefore remove fewer SNPs. When clumping at *r*^2^ = 0.6, the IVW, IVWPCA and F-CLR methods demonstrated a consistently reduced type 2 diabetes mellitus risk with raised *GLP1R* expression, providing further support for this drug indication (Figure 2b and Figure S3; Table S3).

**Figure 2:**
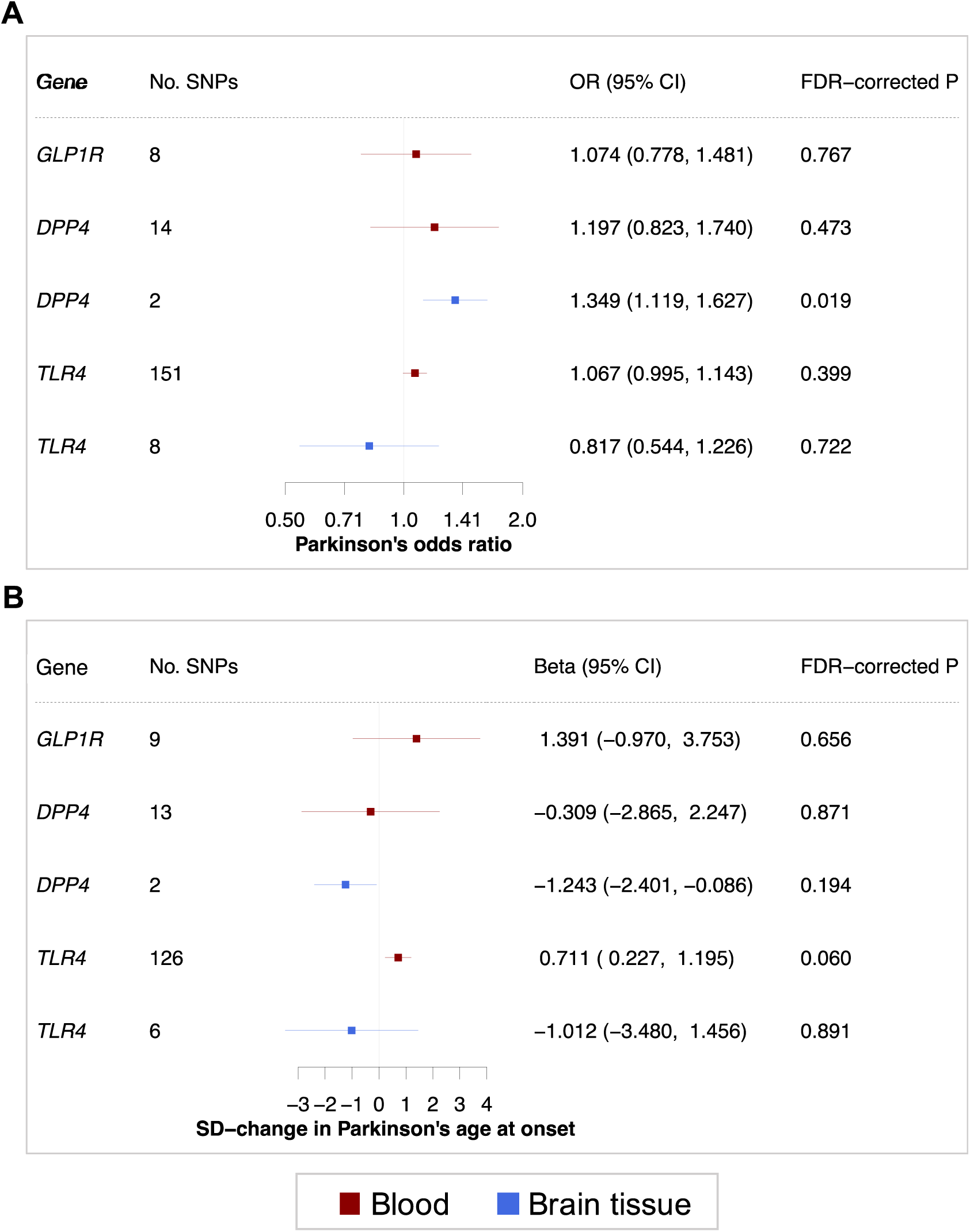
Forest plot illustrating the MR estimates of GLP1R expression in blood, as well as DPP4 and TLR4 expression in in blood and brain tissue. All results computed per 1-SD increase in gene expression. Results are colour-coded according to the tissue (red = blood, blue = brain tissue). P-values were corrected for the number of genes tested using the FDR method. (A) Wald ratio or IVW estimates of GLP1R, DPP4 and TLR4 in blood and brain tissue for Parkinson’s risk, clumping at r^2^ = 0.2. (B) Wald ratio or IVW estimates of GLP1R, DPP4 and TLR4 in blood and brain tissue for Parkinson’s age at onset, clumping at r^2^ = 0.2. 95% CI, 95% confidence interval; OR, odds ratio; SD, standard deviation.

For Parkinson’s, we found no association between *GLP1R* expression in blood and disease risk, age at onset nor any progression outcome (Table S1). Importantly, there were no SNPs associated with *GLP1R* in brain tissue. Raised *DPP4* expression in brain tissue however, which would be associated with reduced brain GLP-1 levels, predicted a significantly raised Parkinson’s risk (Figure 2a and Figure S4; Table S1), and this result passed our quality control (MR-Egger intercept *p* = 0.245, Cochran’s Q *p* = 0.057, *I*^2^ = 0.368). Similarly, greater *DPP4* expression in brain tissue tended to be linked to a younger age at onset, and raised *TLR4* expression in blood was associated with a later age at onset at nominal significance (Figure 2b and Figure S5; Table S2). Although *DPP4* expression in blood was not associated with Parkinson’s risk nor age at onset, the result tended in the same direction.

For the 15 additional genes tested, *AKT3* expression in brain tissue was associated with the Parkinson’s risk and MOCA scores in Parkinson’s, and *GSK3B* expression in blood was associated with developing dyskinesias (Table S1 and S2). Both genes passed the MR-Egger intercept and Cochran’s Q tests for these outcomes. Expression of *AKT1, AKT2, MAPK13* and *MTOR* were associated with BMI (Table S1 and S2).

## Discussion

In this study, we demonstrate that MR using eQTLs can predict the efficacy of a drug; we found that genetically-raised expression of *GLP1R* is causally related to a lower BMI and possibly type 2 diabetes mellitus risk, “rediscovering” the effects of GLP1 receptor agonists in these conditions. While GLP-1 receptor agonists and DPP4 inhibitors are used as symptomatic agents to control blood sugar through effects on insulin release, there is also evidence of a trophic effect on beta islet cells resulting from GLP-1 receptor stimulation that may mitigate the risk of developing type 2 diabetes (Foltynie and Athauda 2020).

We use several MR methods and quality control metrics with different underlying assumptions to probe the robustness of our results, including methods that relax the requirement of strictly independent SNPs. Although exenatide has shown much promise in Parkinsons (Athauda et al. 2017), we found no effect linking peripheral *GLP1R* and Parkinson’s risk, age at onset or progression. Notably, there is previous genetic evidence that a rare variant in *GLP1R* is associated with lower type 2 diabetes mellitus risk but not Parkinson’s risk (Scott et al. 2016).

Moreover, we find that raised *DPP4* expression is associated with an increased Parkinson’s risk. Interestingly, there is longitudinal observational evidence that diabetic patients taking DPP4 inhibitors have a lower incidence of Parkinson’s disease (Brauer et al. 2020). Since DPP4 breaks down GLP-1, this indicates that any protective actions of GLP-1’s may not involve GLP1R in blood and that exenatide may be effective in Parkinson’s through an alternative mechanism.

It is unclear whether any effects of GLP-1 receptor agonists in Parkinson’s are related to peripheral or central GLP1R stimulation. We found no eQTLs for *GLP1R* in brain tissue, and Parkinson’s risk, age of onset or progression may be modulated by GLP1R stimulation in the brain. This explanation is supported by our results that raised *DPP4* and reduced *TLR4* expression in brain tissue may be linked to a younger age at onset of Parkinson’s. Although these genes reached nominal significance for age at onset, this trend further suggests that GLP-1 may influence Parkinson’s independently of GLP1R in blood. Furthermore, Athauda and colleagues analysed the neuronal-derived exosomes from Parkinson’s patients in the Exenatide-Parkinson’s trial, and they found that patients treated with exenatide had elevated total Akt at 48 weeks (Athauda et al. 2019). When looking at 15 additional proteins thought to be involved in the exenatide pathway, we found evidence for target engagement with the Akt-signalling pathway. This potently illustrates how MR can be used to explore molecular mechanisms of action.

Although we have included the largest Parkinson’s progression GWAS available, there is a possibility that exenatide acting on *GLP1R* in blood has a weaker effect on Parkinson’s progression than is detectable by this MR study. For our disease risk and age at onset outcomes, our power is boosted by large GWAS sample sizes. This MR study therefore mostly pertains to whether exenatide could prevent or delay disease, rather than halt progression. This is an important consideration, because previous work suggests a disconnect between the molecular mechanisms driving Parkinson’s risk versus progression (Storm et al. 2020; Nalls et al. 2019; Iwaki et al. 2019; Blauwendraat et al. 2019).

Furthermore, increased *GLP1R* expression in blood may not accurately represent the biological consequences of exenatide, which involve GLP-1 receptor stimulation in pancreatic cells. It may be more appropriate to use expression data from biologically relevant tissue such as the pancreas for diabetes and the brain for Parkinson’s disease, however the sample sizes of current tissue-diverse eQTL datasets are small compared to whole-blood projects. Similarly, SNPs associated with protein levels (pQTLs) may be a more suitable mimic, however to our knowledge no pQTL has been found for the GLP-1 receptor.

While the randomized controlled trial remains the gold-standard for evaluate a drug, MR has shown promise in predicting the success of a drug. Two MR studies about the effect of serum urate levels on Parkinson’s found no causal effect (Kia et al. 2018; Kobylecki and Nordestgaard 2018), and sooner thereafter a phase III clinical trial was terminated ahead of schedule due to insufficient efficacy (https://www.ninds.nih.gov/Disorders/Clinical-Trials/Study-Urate-Elevation-Parkinsons-Disease-Phase-3-SURE-PD3/). Many advocate the use of MR and QTL data in in drug development (Evans and Davey Smith 2015; Schmidt et al. 2020; Storm et al. 2020), and this project provides a valuable example for the potential and limitations of this approach.

## Data Availability

The GWAS data used by this study are publicly available as stated in the original publications. The supplementary information contains full results. We make our code openly available at https://github.com/catherinestorm/mr_exenatide.

## Contributors

CSS, DAK, MA, NWW contributed to the idea, design, interpretation and verification of the study. CSS performed the analyses and drafted the manuscript, with input from all authors. SB provided advice on the methods used in this study. DA and TF contributed to the intepretation of these results and the genes studied. This project is part of ongoing work by the IPDGC, and all Parkinson’s disease GWAS data used here was curated and made available by members of the IPDGC. All authors critically revised and commented on the manuscript before submission.

## Acknowledgements

CSS would like to thank Vishal Rawji for his continued encouragement, support and outside perspective throughout the production of this study. We thank Ashish Patel for kindly sharing the code used for the F-LIML and F-CLR methods. CSS is funded by Rosetrees Trust, John Black Charitable Foundation and the University College London MBPhD Programme. DAK is supported by an MBPhD Award from the International Journal of Experimental Pathology. MA is funded by the Faculty of Applied Medical Sciences, King Abdulaziz University, Jeddah, Saudi Arabia. NWW is a National Institute for Health Research senior investigator and receives support from the European Union Joint Programme— Neurodegenerative Disease Research Medical Research Council Comprehensive Unbiased Risk factor Assessment for Genetics and Environment in Parkinson’s disease. NWW receives support from the National Institute for Health Research University College London Hospitals Biomedical Research Centre. We thank the members of the IPDGC and authors of the referenced QTL projects for making the their GWAS data openly available. Finally, we thank all the patients and families whose decision to donate tissue samples to genetic research made our project possible.

## Supplementary material

Table S1: Full MR results for all genes tested.

Table S2: MR quality control (MR Egger intercept, Cochran’s Q, *I*2 tests) for all genes tested.

Table S3: Results for *GLP1R* from IVW, IVWPCA, FLIML and FCLR methods when clumping at r^2^ = 0.6.

Supplementary Note: Full list of IPDGC members and affiliations.

**Figure S1:**
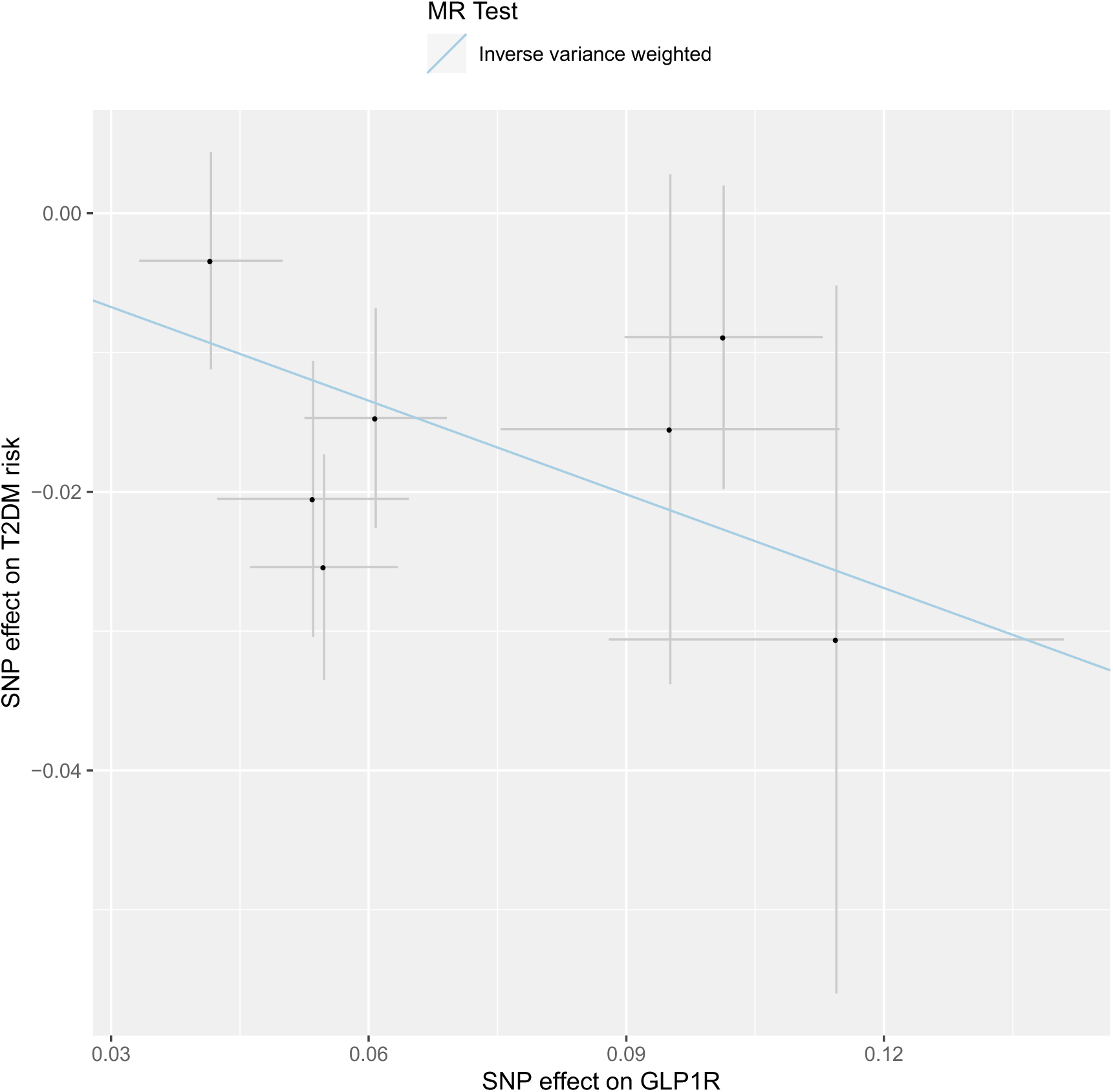
Scatter Plot. GLP1R and type 2 diabetes type 2 diabetes; blood.

**Figure S2:**
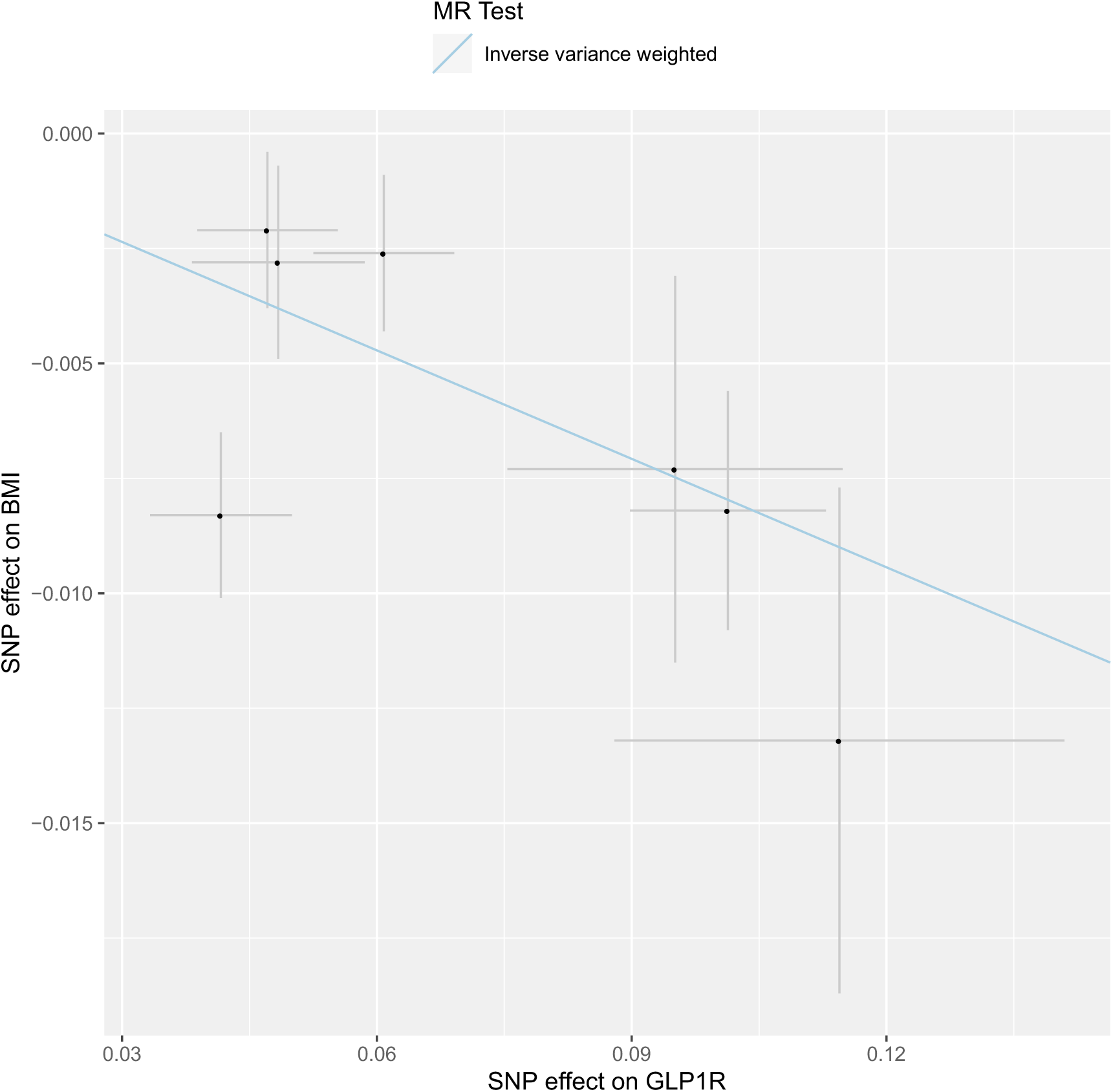
Scatter Plot. GLP1R and BMI; blood.

**Figure S3:**
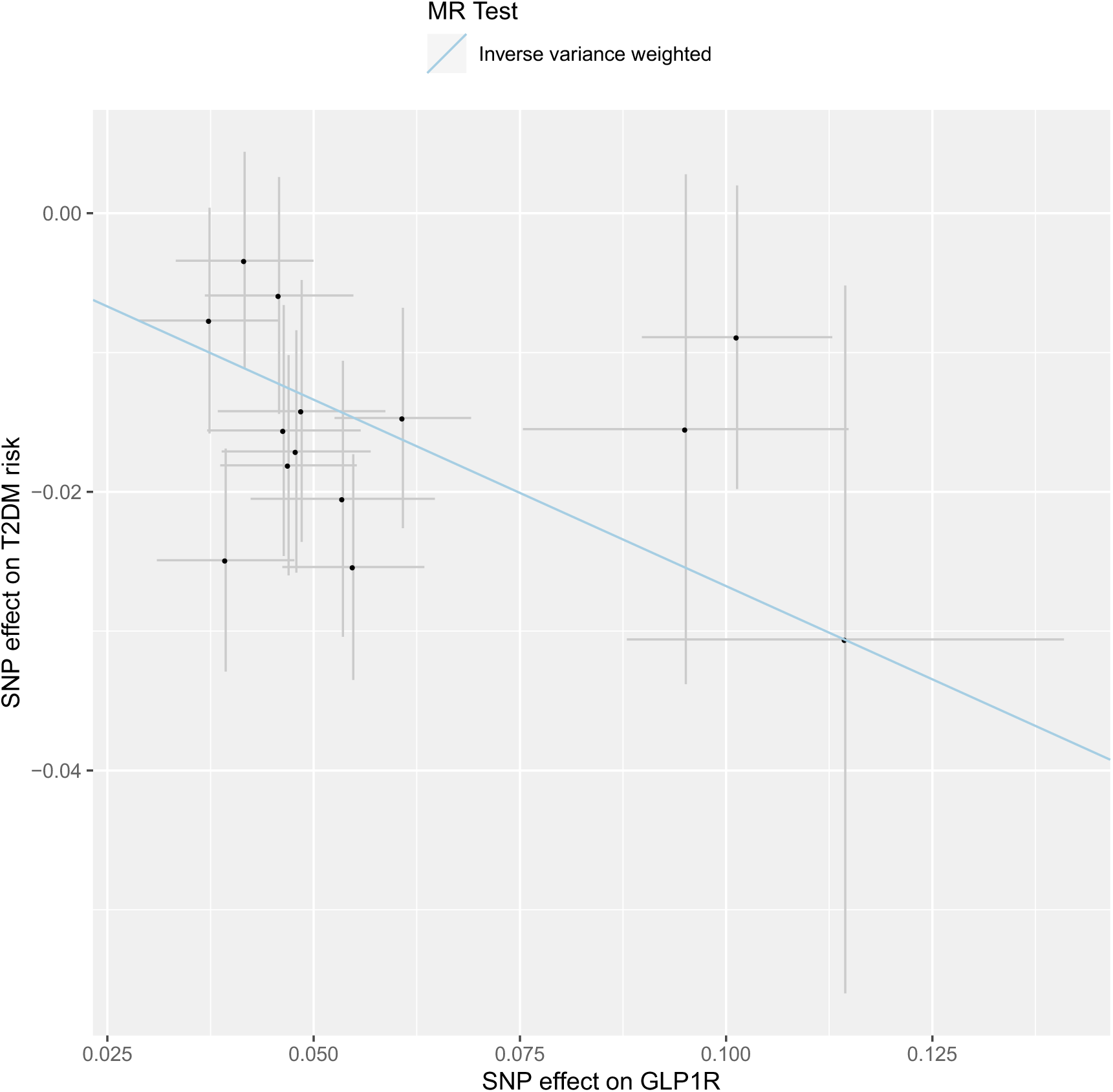
Scatter Plot. GLP1R and type 2 diabetes risk clumping at r^2^ = 0.6; blood.

**Figure S4:**
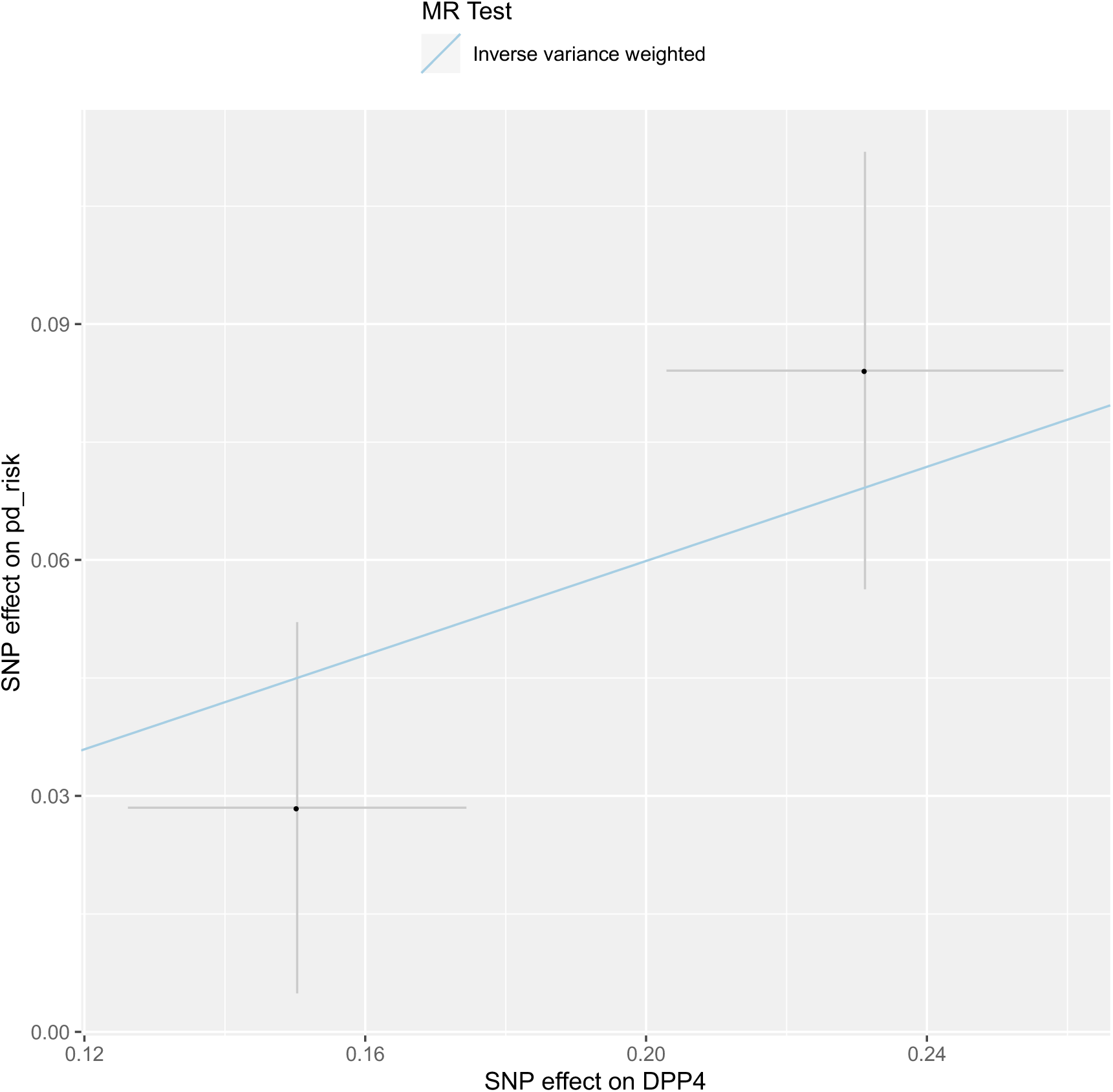
Scatter Plot. DPP4 and Parkinson’s disease risk; brain tissue

**Figure S5:**
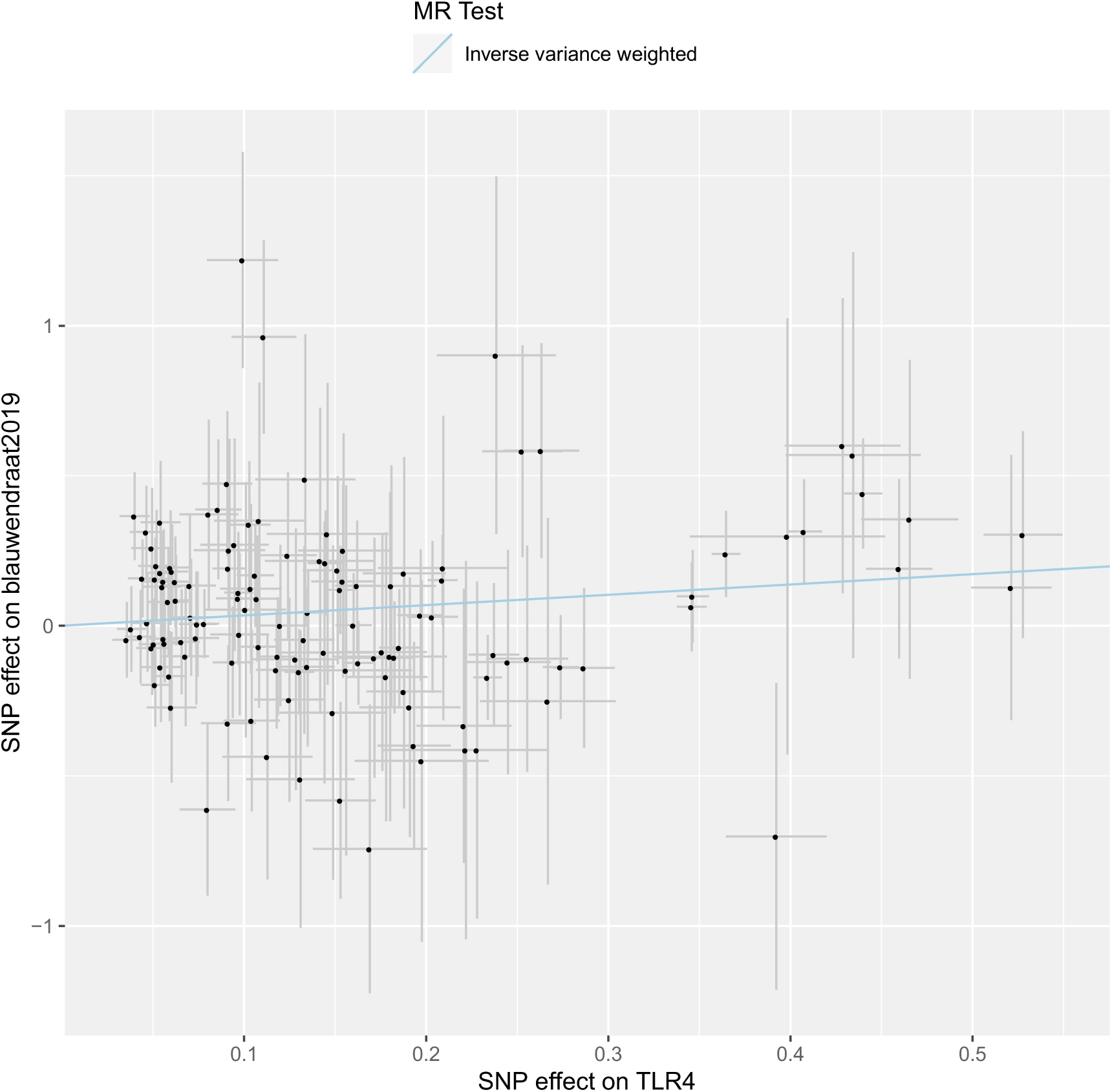
Scatter Plot. TLR4 and Parkinson’s disease age at onset; blood.

